# Covid-19 epidemic curve in Brazil: A sum of multiple epidemics, whose income inequality and population density in the states are correlated with growth rate and daily acceleration

**DOI:** 10.1101/2020.09.09.20191353

**Authors:** Airandes de Sousa Pinto, Carlos Alberto Rodrigues, Carlito Lopes Nascimento Sobrinho, Edval Gomes dos Santos, Lívia Almeida da Cruz, Paulo Cesar Nunes, Matheus Gomes Reis Costa, Manoel Otávio da Costa Rocha

## Abstract

**Introduction:** Recently, we demonstrated that the polynomial interpolation method can be used to accurately calculate the daily acceleration of cases and deaths by Covid-19. The acceleration of new cases is important for the characterization and comparison of epidemic curves. The objective of this work is to measure the diversity of epidemic curves and understand the importance of socioeconomic variables in the acceleration, peak cases and deaths by Covid-19 in Brazilian states.

**Methods:** This is an ecological study with time series analysis of new cases and deaths by Covid-19 in Brazil and its 27 federation units. Using the polynomial interpolation method, we calculated the daily cases and deaths with the measurement of the respective acceleration. We calculated the correlation coefficient between the epidemic curve data and socioeconomic data.

**Results:** The combination of daily data and acceleration determined that the states of Brazil are in different stages of the epidemic. Maximum acceleration of peak cases, peak of cases, maximum acceleration of deaths and peak of deaths are associated with the Gini index and population density, but did not correlate with HDI and per capita income.

**Conclusion:** Brazilian states showed heterogeneous data curves. Density population and socioeconomic inequality are associated with worse control of the epidemic.

## INTRODUCTION

Brazil registered the first case of Covid-19 on 02/26/2020 in São Paulo (1). The country developed one of the highest growth rates for new cases of Covid-19 in the world (2), reaching, on July 11, 1,839,850 cases and 71,469 deaths. However, the epidemic curve does not present a homogeneous behavior in the population, and a regional and ethnic variation in mortality by Covid-19 in Brazil has been described (3).

The epidemic curve has been obtained, routinely, by the 7-day moving average of the events. Although it facilitates the visualization of categorical variables, which present a large daily variation, it does not allow the calculation of the absolute daily variation rate, that is, daily acceleration. Recently, we demonstrated that the polynomial interpolation method can be used to accurately calculate the daily acceleration of cases and deaths by Covid-19 (4). This method, which applies a methodology that uses the derivative of differential calculus, is used to study curves in several areas of knowledge (5–7).

The measurement of the acceleration contributes to the classification of the epidemic curve, comparison of curves between countries and states, in addition to assisting in the application of non-pharmacological measures to face the Covid-19 epidemic. This variable is responsible for the slope of the curve in the ascending and descending phases of the spread and its lower absolute value in the first phase may indicate the adoption, by countries and states, of more appropriate measures to combat the pandemic, corresponding to a flattended curve in the graph.

Brazil has a high level of socioeconomic inequality that can impact the intensity of acceleration and peak of daily cases and deaths. The objective of this work is to measure the diversity of epidemic curves and understand the importance of socioeconomic variables in the acceleration, peak cases and deaths by Covid-19 in Brazilian states.

## METHODS

### Study design and population

This is an ecological study with time series analysis of new cases and deaths by Covid-19 in Brazil and its federation units.

We studied the 27 federation units: Acre, Alagoas, Amapá, Amazonas, Bahia, Ceará, Espírito Santo, Goiás, Maranhão, Mato Grosso, Mato Grosso do Sul, Minas Gerais, Pará, Paraíba, Paraná, Pernambuco, Piauí, Rio de Janeiro, Rio Grande do Sul, Rio Grande do Norte, Rondônia, Roraima, Santa Catarina, São Paulo, Sergipe, Tocantins and Distrito Federal (the Federal District). The time series for each unit of the federation were obtained from the website of the Brazilian Ministry of Health from February 25 to July 11 (0 to 137 of the historical series) (8). The Gini index (developed by the Italian mathematician Conrado Gini in 1912, is used to measure the distribution of income across a social group. It points out the difference between the income of the poorest and the richest in a population, numerically varies from zero, 0, perfect equality to one, 1, perfect inequality) (9), per capita income (ratio between the total income obtained and the total number of inhabitants in a given state/territory - numerical indicator), population density (ratio between the total population and a given territory (geographical area) - numerical indicator)(10), HDI (Human Development Index-developed in 1990, by economists Nahbub ul Haq (Pakistani) and Amartya Sem (Indian), it is a combination of life expectancy, average schooling and per capita income of a given population, numerically ranges from zero (0) very low HDI one (1) very high HDI) (9). The HDI of the population of Brazil and federation units was obtained from the IBGE website.

We calculated the incidence rate by the ratio between the number of cases accumulated on the last day of the series and the population multiplied by 1000. Likewise, mortality was obtained by dividing the number of deaths accumulated at the end of the historical series and the population multiplied by 1000.

### Mathematical modeling

The epidemic curve was obtained by using the polynomial method in a similar way in previous work (4).

The polynomial was automatically generated by the Matlab software, whose degree and coefficients were adjusted to the curve of daily cases and deaths, figure 1, using the degree at most 8, according to the equation

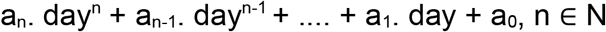

**Figure 1.**
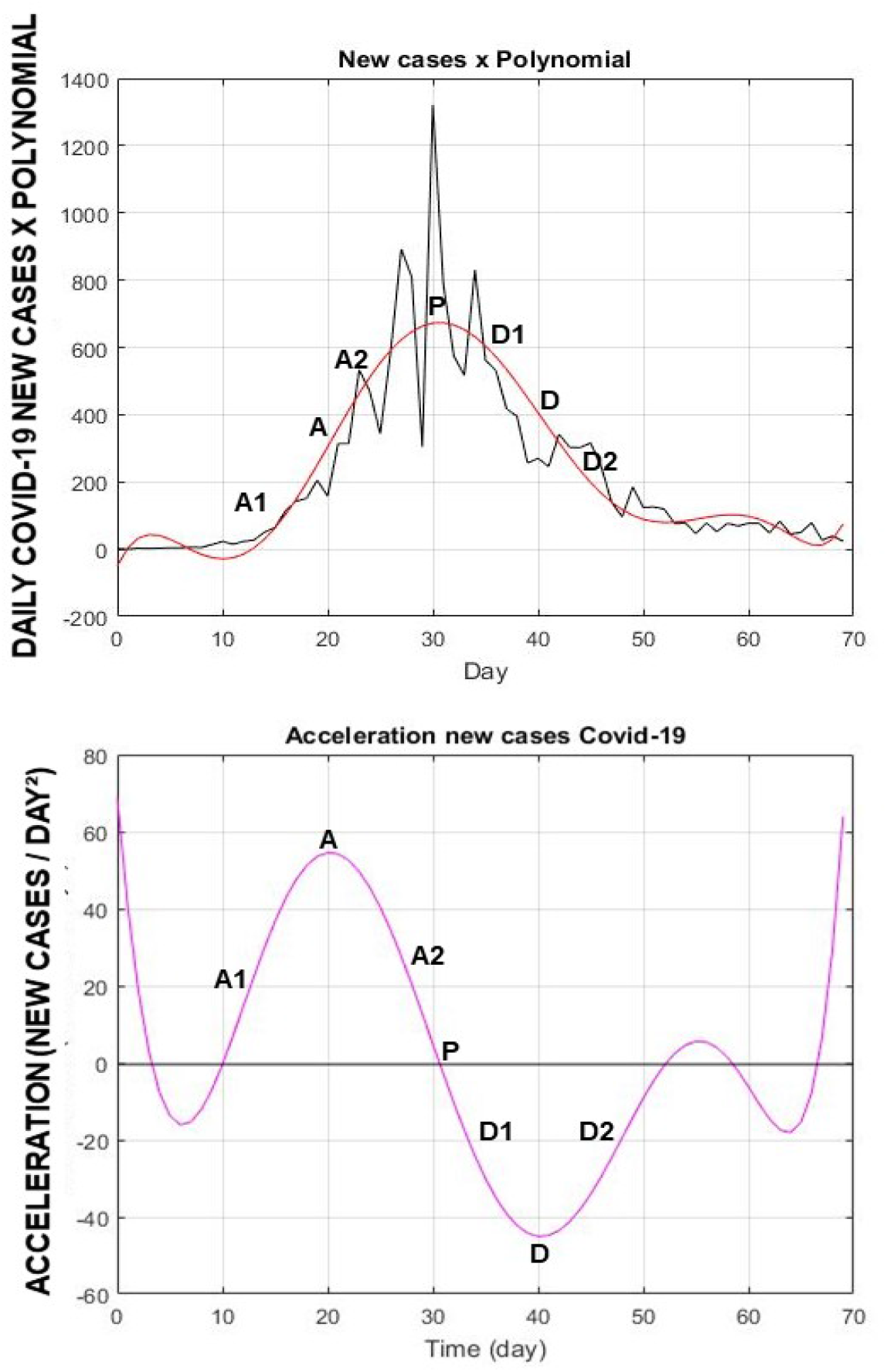
Upper: Daily cases of Covid-19 from Austria x polynomial. Lower: daily acceleration of Covid-19 from Austria.. A: Maximum acceleration. A1: first stage of the ascending phase, acceleration increases to maximum value, A2: acceleration remains positive, but decreases value to zero. P: Peak daily cases obtained by the polynomial. D: Absolute maximum value of the descending phase of new cases. D1: First stage of the descending phase, absolute values increase to maximum value, D. D2: second stage of the decreasing curve, deceleration values are reduced to zero.

In order to illustrate the applicability of the polynomial in describing the epidemic curve, we present data on new cases from Austria, a country that presented the complete curve at the end of this historical series, **figure 1**.

The instantaneous acceleration was obtained by the first derivative of the polynomial. The day of the maximum acceleration value and the day of the maximum deceleration value, when present, were determined by the second derivative of the polynomial. In the descending phase of the curve, the negative values of the acceleration were analyzed taking into account their absolute values and reported as deceleration, **figure 1**. In the states where daily acceleration was still increasing until the end of the series, the instantaneous acceleration was evaluated up to five days before day 137, the end date of the series, to avoid instability in the polynomial that is observed at the end of the acceleration curve.

The peak day of new infections was evaluated by the root of the polynomial.

The epidemic curve of each state was classified according to the values of cases and acceleration. The Covid-19 infection graph was used to classify the epidemic into phases: ascending, peak and descending. The acceleration graph made it possible to subdivide these phases. The ascending phase presents a first stage with a concomitant increase in new infections and acceleration and a second stage in which numbers continue to increase, but there is a gradual decrease in acceleration. At the peak, the number of new cases stabilizes and the acceleration reaches zero. The descending phase also has two stages. In the first stage, there is a decrease in new cases and an increase in the absolute value of the deceleration. In the second stage, the event value continues to decrease, but the acceleration returns to zero, **figure 1**.

### Statistical analysis

The correlations of socioeconomic data, incidence rate, mortality, maximum acceleration of cases, peak of cases and deaths were measured by the Pearson Correlation Coefficient if they presented normal or normalized distribution after logarithmic transformation. The Spearman method was used to assess the correlation between variables with non-normal distribution.

The normality analysis was performed by the Kolmogorov-Smirnov tests.

The results were considered significant if p <0.05.

## RESULTS

### Cases

On July 11, 2020, the last day of the historical series, Brazil reached 1,839,850 cases, incidence rate = 8.94 cases/1000 inhabitants, table 1. Brazil presented the maximum acceleration = 604.7 cases/day^2^ on May 21st, 86th day of the series. The most recent acceleration is equal to 373.75 new cases/day^2^, showing a reduction, but still remaining high. As it presented a decrease in acceleration, the country was in the second stage of the acceleration phase, **figure 2**.

**Table 1.**
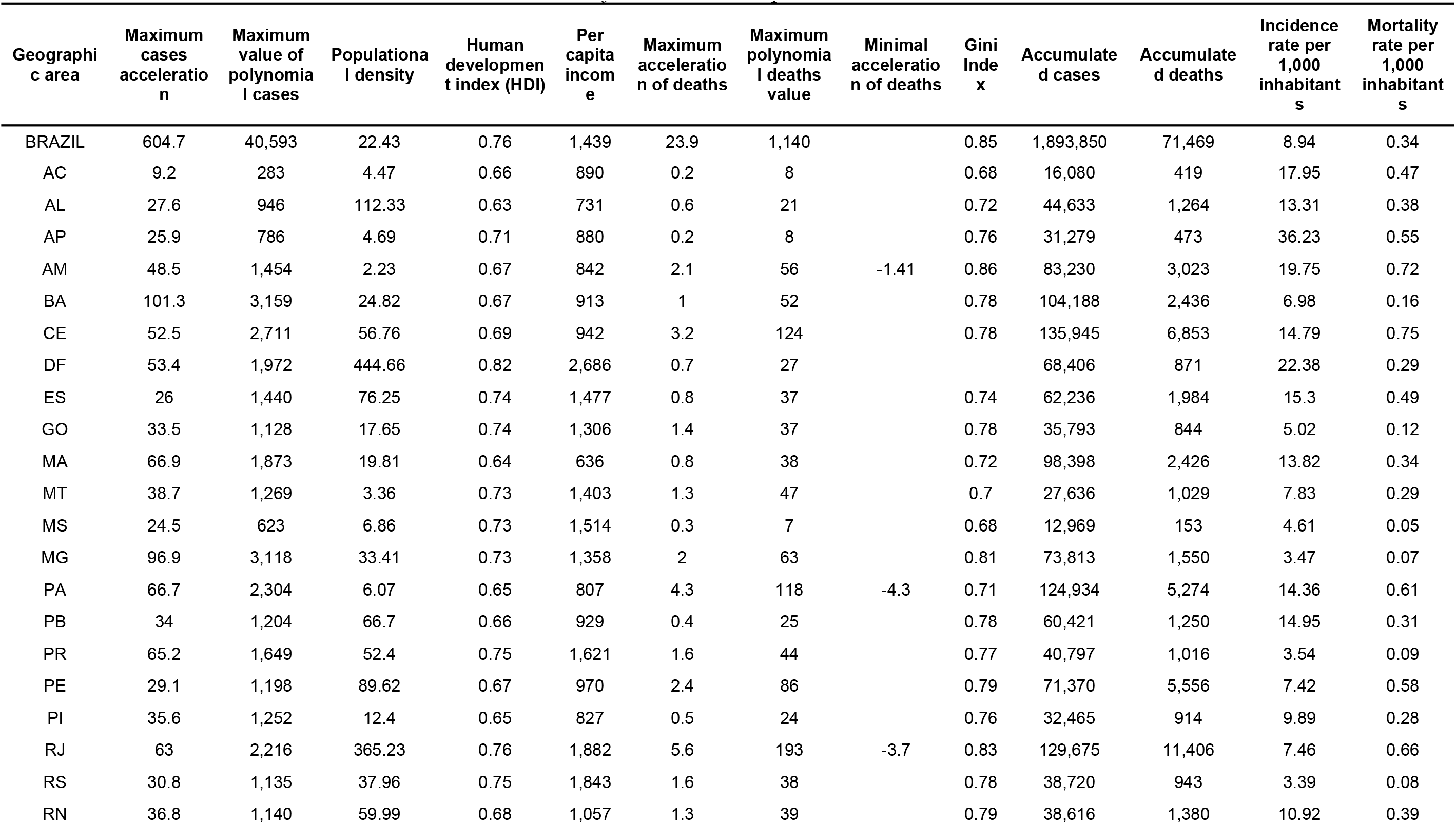
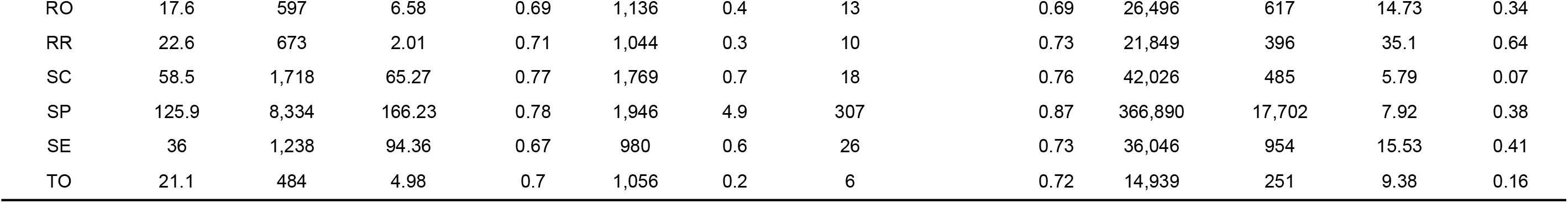
*Socioeconomic variables and variables obtained from Covid-19 epidemic curves*.

**Figure 2.**
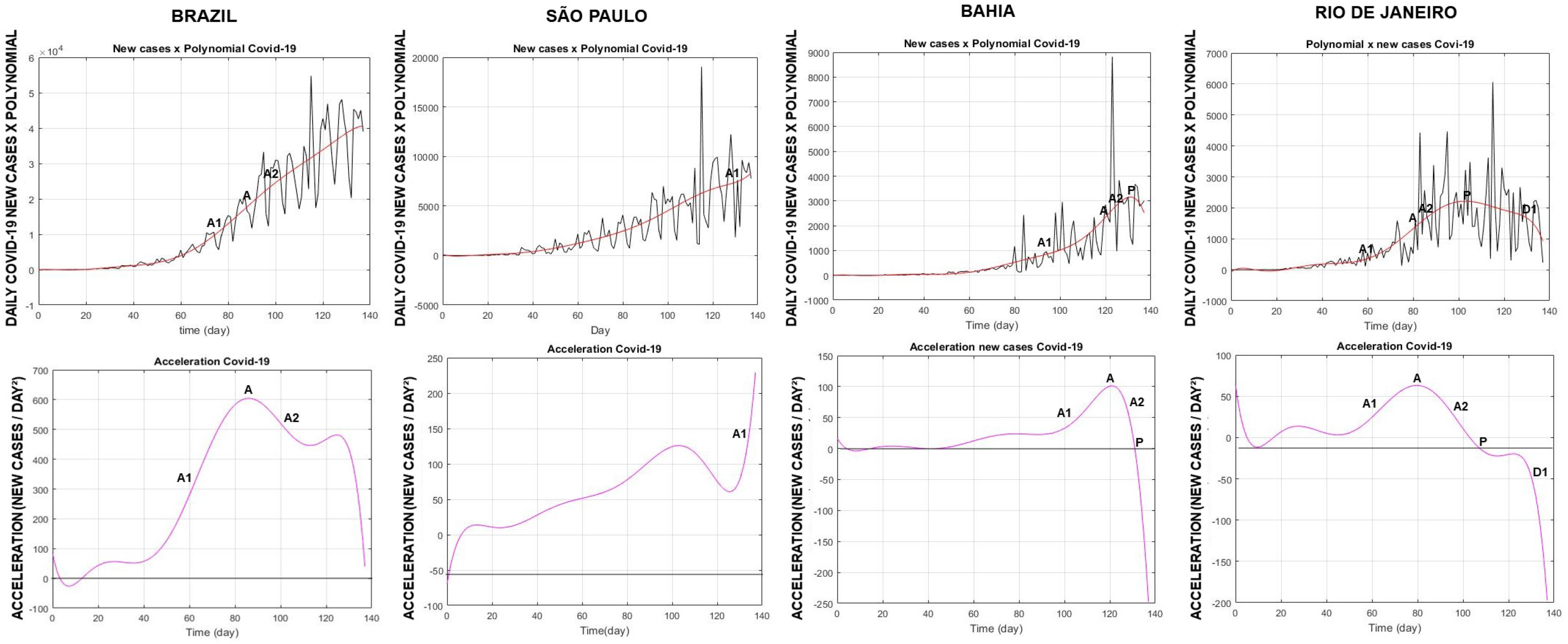
Upper: New cases of Covid-19 in Brazil, São Paulo, Bahia and Rio de Janeiro x polynomial. Lower: daily acceleration of new cases of Covid-19 in Brasl, São Paulo, Bahia and Rio de Janeiro. A: Maximum daily acceleration. A1: first stage of the growth phase. A2: second stage of the growth phase. P: acceleration equal to zero (peak). D1: first stage of the descendant phase.

The peak of new cases showed a correlation with population density, r = 0.53, p = 0.004 and with the Gini index, r = 0.49, p = 0.01, but there was no significant correlation with the HDI or with per capita income. The maximum daily acceleration of cases correlated with population density, r=0.43,p=0.03 and with the Gini index, r=0.45,p=0.02, **table 2**.

**Table 2.** Correlation between socioeconomic variables and variables of the Covid-19 curve in Brazilian states.

|  | Maximum cases acceleration | Peak of cases | Maximum acceleration of deaths | Peak of deaths | Incidence rate per 1,000 inhabitants | Mortality rate per 1,000 inhabitants |
| --- | --- | --- | --- | --- | --- | --- |
| **Gini Index** | r =0.45<br>p= 0.02 | r=0.49<br>p= 0.01 | rs =0.64 *<br>p<0.001 | rs= 0.62<br>p= 0.001 | r= -0.31<br>p= 0.12 | r = 0.18<br>p= 0.38 |
| **HDI** | r=0.25<br>p= 0.22 | r=0.28<br>p= 0.16 | r=0.17<br>p= 0.39 | r= 0.14<br>p= 0.49 | r= -0.31<br>p= 0.11 | r= -0.30<br>p= 0,12 |
| **Per capita income** | r= 0.24<br>p= 0.23 | r= 0.28<br>p= 0.15 | r=0.22<br>p= 0.27 | r= 0.19<br>p= 0.35 | rs= -0,42<br>p= 0,03 | rs= -0.38<br>p=0.053 |
| **Populational density** | r=0.43<br>p= 0.03 | r=0.53<br>p= 0.004 | r=0.40<br>p=0.04 | r=0.50<br>p= 0.009 | r=-0,24<br>p= 0,22 | rs= -0.02<br>p= 0.91 |

The highest incidence rate occurred in Amapá, followed by Roraima and Distrito Federal; 36.23 cases/1000, 35.10 cases/1000 and 22.38 cases/1000, respectively. The lowest incidence rate was observed in Rio Grande do Sul, 3.39 cases/1000.

São Paulo had the highest rate of acceleration, 125.9 cases/day^2^ and the lowest rate of acceleration was observed in Acre with 9.2 cases/day^2^. São Paulo had the highest number of new cases, 8,334 new cases/day and Acre the lowest, 283 new cases/day.

We identified 14 states in the rising phase of the curve. Piauí, Sergipe, Mato Grosso, Mato Grosso do Sul, Minas Gerais, São Paulo (figure 2), Santa Catarina and Rio Grande do Sul are in the first stage of the growth phase, numbers continue to increase concomitantly with the rise in acceleration. Other states like Paraiba, Tocantins, Ceará and Pernambuco had a peak of new infections, but the number of cases rose again, surpassing the previous number, and are classified in the ascending phase of the epidemic curve. Goiás and Paraná are in the second stage of the ascending phase

The states of Acre, Amazonas, Pará, Rondônia, Roraima, Alagoas, Bahia (figure 2), Maranhão and Espírito Santo showed stable behavior, being classified in the peak phase of the epidemic curve.

Amapá, Rio Grande do Norte, Distrito Federal and Rio de Janeiro (figure 2) presented consistent negative acceleration, classified in the first stage of the deceleration curve, new cases decreasing with the deceleration, however, increasing, without reaching yet the maximum value. Rio de Janeiro has the highest deceleration value, -71.7 cases/day^2^, day 132th day of the serie, **figure 2**.

### Deaths

On July 11, 2020, the last day of the historical series, Brazil registered 71,469 deaths, mortality equal to 0.34 deaths/1000 inhabitants. The country presented the maximum acceleration = 23.9 deaths/day^2^ on May 7th, day 83 of the series. The most recent acceleration is equal to 13.4 deaths/day^2^, after a short period in which it was negative, constituting a peak in plateau, **table 1 and figure 3**.

**Figure 3.**
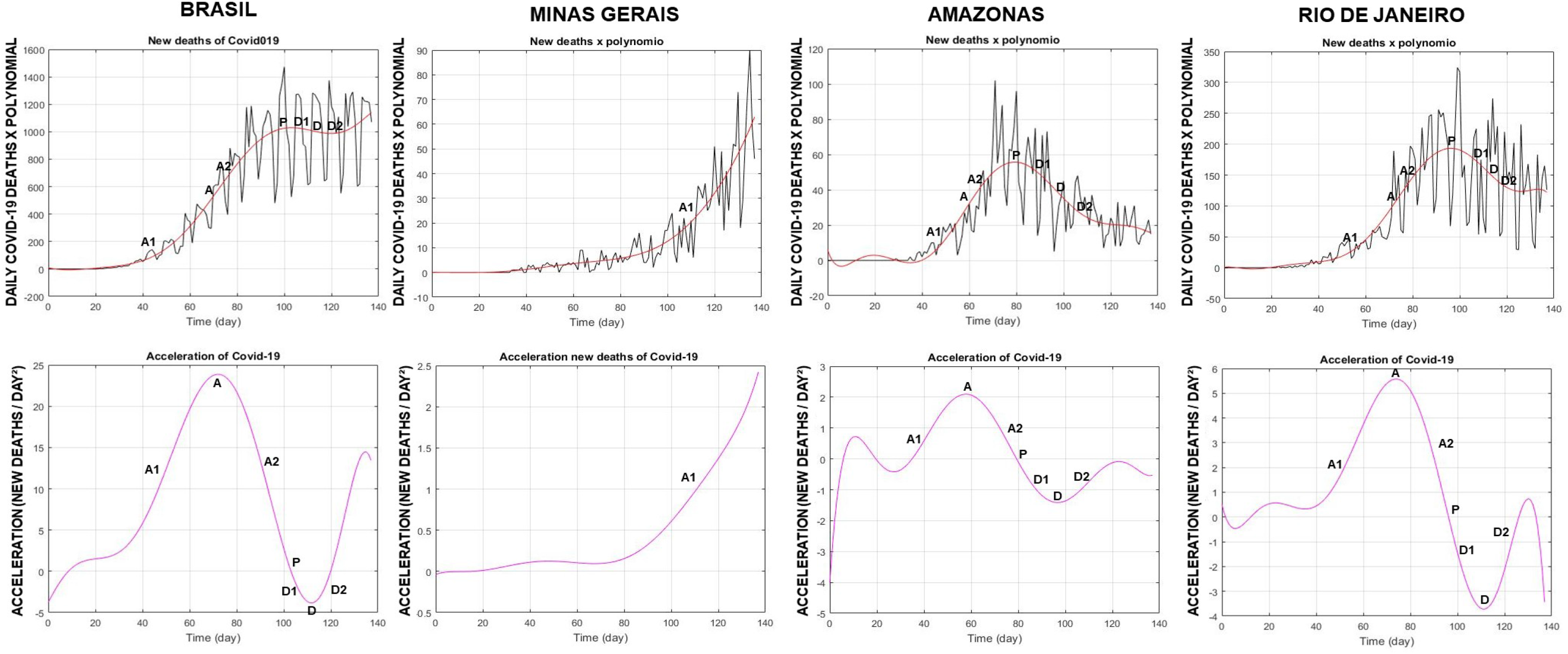
Upper: Daily deaths by Covid-19 in Brazil, Minas Gerais, Amazonas and Rio de Janeiro x polynomial. Lower: daily acceleration of deaths in Brazil, Minas Gerais, Amazonas and Rio de Janeiro. A: Maximum daily acceleration. A1: first stage of the growth phase. A2: second stage of the growth phase. P: acceleration equal to zero (peak). D1: first stage of the decline phase.

The maximum value of deaths was correlated with population density, r = 0.50, p = 0.009 and with the Gini index, rs = 0.62, p =0.001. The maximum acceleration of deaths was correlated with population density, r = 0.40, p = 0.04 and with the Gini index rs = 0.64, p <0.001. The HDI and per capita income did not show any significant correlation with the maximum acceleration of daily deaths or with the maximum value of daily deaths, **table 2**.

The incidence rate / 1000 inhabitants showed a correlation with per capita income, rs = -0.42 p = 0.003; without correlation with HDI, Gini index or population density. The mortality rate / 1000 inhabitants showed a tendency towards a weak correlation with per capita income, rs=-0,38 p=0,053 and no correlation with Gini index, HDI or population density**, table 2**.

The states of Ceará, Amazonas and Rio de Janeiro had the highest mortality rate, 0.75 deaths/1000, 0.72 deaths/1000 and 0.66 deaths/1000, respectively. The lowest mortality rate occurred in Mato Grosso do Sul, 0.05 deaths/1000, **table 1**.

The state of Rio de Janeiro had the highest acceleration rate of deaths, 5.6 deaths/day^2^ and Tocantins the lowest acceleration, 0.2 deaths/day^2^. São Paulo had the highest number of deaths/day assessed by the polynomial, 307 deaths/ day. The lowest value occurred in Tocantins with 06 deaths/day.

The states of Rio Grande do Sul, Santa Catarina, Paraná, Minas Gerais (figure 3) and Mato Grosso are in the first stage of the growth phase of the curve. There are increases in cases accompanied by increased daily acceleration. The states of Mato Grosso do Sul, Goiás, Distrito Federal, Paraíba and Bahia were in the second stage of the growth phase of the death curve, an increase in deaths/day associated with positive acceleration, but with a decrease in the value. São Paulo and Tocantins peaked, but the death curve rose again, therefore, in a new phase of acceleration. Ceará was in the descending phase of the curve, but accelerated again.

Sergipe and Roraima showed a peak behavior at the end of the historical series. Alagoas, Pernambuco, Maranhão and Rondônia had a peak in plateau.

Rio de Janeiro, **figure 3**, Espírito Santo, Rio Grande do Norte, Piauí, Amapá, Acre were at the end of the first stage of degrowth. Amazonas, figure 3, and Pará were in the second stage of the descending phase.

## DISCUSSION

Our results demonstrate that the peak of daily cases, acceleration of cases and deaths and peak of deaths showed a positive correlation with population density and the Gini Index. This result is in agreement with articles that demonstrate the importance of population distancing for the flattening of the epidemic curve, and reinforce the usefulness of lockdown or physical distancing to reduce peak cases and deaths (11).

Brazil presented a great acceleration of new cases, and, at the end of this series, it was approaching the peak. This curve represents the sum of various epidemics already occurring in the states, in different phases and at different severity levels. Such heterogeneity may be related to the central government’s lack of coordination regarding measures to control the spread of the virus, territorial extent and the country’s geographical, social, economic and cultural diversity. In addition, unlike other epidemic diseases, the Covid-19 epidemic in Brazil had its first reports of imported cases (aloctenes), brought by subjects who returned from international trips or foreigners from countries that were themselves in epidemic situation, and later extended to the periphery and smaller cities in the interior of the country (8).

The results of this study suggest that the worst Covid-19 indicators were related to demographic density and the Gini Index (indicator of income inequality), same behavior observed with social problems that are more related to income inequality than per capita income (12).

The incidence and mortality rate data should be viewed with caution, as we are still experiencing the epidemic and, at this moment, we need data that reflect the daily variation of newly reported cases and their acceleration, which are important because they reflect the strength or trend of the outbreaks taking daily data into account. It does not need to be compared with previous data, it assesses the absolute slope of the curve. Its measurement can be useful to evaluate non-pharmacological measures in response to the Covid-19 pandemic.

## CONCLUSION

Brazilian states presented heterogeneous data curves, therefore classified at different stages of the epidemic, with socioeconomic inequality and population density being associated with worse control of the epidemic.

## Data Availability

All data were obtained from public websites of the government of Brazil.

https://www.ibge.gov.br

https://www.saude.gov.br

